# Electoral outcomes and health inequality – association between respiratory disease and voting patterns in England

**DOI:** 10.1101/2025.06.04.25327439

**Authors:** Anthony A. Laverty, Nicholas S Hopkinson

**Affiliations:** School of Public Health, Imperial College London; National Heart and Lung Institute, Imperial College London, London, UK

## Abstract

**Objectives:** To establish if there is an association between health metrics and patterns of voting in England, particularly in relation to a recently established right wing party, Reform UK, in the 2024 general election.

**Design:** Ecological cross-sectional study with linear regression

**Setting:** England

**Participants:** 543 constituencies in England

**Main outcome measures:** Estimated prevalence of 20 common non-communicable diseases, including obesity, COPD, asthma, type 2 diabetes and depression.

**Results:** Constituencies electing Reform MPs (n=5/543) had the highest prevalence of Asthma (7.44%) and COPD (2.85%). Across the country, adjusting for age, sex and deprivation, a 10% increase in the party’s vote share was associated with a +0.261% (95% Confidence Interval 0.213 to 0.309) prevalence of COPD, a +0.113% (95%CI 0.026 to 0.201) prevalence of asthma and a +1.479% (95%CI 1.239 to 1.720) increase in obesity prevalence.

**Conclusions:** At a constituency level, poor health, in particular conditions associated with breathlessness, is associated with a greater proportion of votes for Reform UK.

## INTRODUCTION

Health is a fundamental issue in politics - essential both in relation to the well-being of citizens and for a nation’s economic productivity. Decisions by policy makers influence both the provision of healthcare services and the upstream social and commercial determinants of health including clean air, transport, food, housing and employment. Lung health is particularly influenced by health inequality[1-4]. At the same time, experience of good or poor health and interactions they have with the healthcare system may influence voters’ perceptions of incumbent political parties and the alternatives being offered to them. It is therefore to be expected that there will be interactions between health and public opinion, in particular, the way that opinion is expressed through voting in elections.

In the July 2024 UK general election, Reform UK, a political party founded in 2018, secured 14% of votes, winning 5 out of the 543 English constituencies and therefore 5 Members of the UK Parliament. More recently, in May 2025, the party has won a substantial number of council seats in local authority elections. Many countries in Europe and elsewhere have seen a rise in populist right-wing parties and there is evidence that support for them is linked to health and satisfaction with healthcare services[5-9]

We therefore wished to establish whether there was a relation between constituency level health metrics in England and voting patterns, particularly in relation to this new party.

## METHODS

Data from the 2024 General Election, including size of electorate, number of valid votes and votes per political party came from the House of Commons Library[10]. Our analyses are all at the constituency level for England only. We used two measures of the strength of support for Reform UK: 1) constituencies in England returning a Reform MP compared to those electing a different political party 2) the proportion of votes for Reform across all constituencies.

Health outcomes were the prevalence of 20 common health conditions collated by the House of Commons Library based on the NHS Quality and Outcomes Framework data from 2022/23: asthma; atrial fibrillation; cancer diagnosis (any); chronic kidney disease (CKD); COPD; coronary heart disease (CHD); dementia; depression; type 2 diabetes; epilepsy; heart failure; hypertension; learning disabilities; non-diabetic hyperglycaemia; obesity; osteoporosis; peripheral arterial disease; rheumatoid arthritis; schizophrenia, bipolar disorder and psychoses; and stroke/TIA (combined)[11]. We took age and sex data of resident populations also from the Office for National Statistics population estimates for 2022 as well as Index of Multiple Deprivation (IMD) data which compares constituencies on their relative deprivation; divided into five groups for analyses[11 12].

We first compared prevalences of each health condition according to the MP returned in the General Election (Labour, the Conservative and Unionist Party, the Liberal Democrats, the Green Party and Reform UK). We used Pearson Correlation Coefficients to investigate relationships between Reform UK vote share and health metrics. Finally, we conducted separate linear regression models for each health metric, controlling for age, sex and deprivation. We did not involve patients or the public in this paper.

## RESULTS

Of 543 constituencies in England, Labour won 347, the Conservatives 116, the Liberal Democrats 65, the Green Party 4 and Reform UK won 5 seats (total = 537) in the 2024 General Election. We excluded data from constituencies returning an independent MP (n=5), as well as the constituency of the Speaker of the House (n=1), as by convention the main parties did not put up a candidate against him.

Of the five areas returning a Reform UK MP, 3 (60%) were the most deprived fifth of the country (**Table 1**). 103 (29.7%) Labour voting areas were the most deprived. Reform areas had the highest proportions of residents aged over 65 years of age (23.8% vs. 17.1% for Labour and 23.2% the Conservatives).

**Table 1:** Age, sex and deprivation breakdown of constituencies by winning party in 2024 UK General Election.

| **Winning party 2024** | **N** | **N (%) most deprived fifth\*** | **Mean % over 65** | **SD** | **Mean % Male** | **SD** |
| --- | --- | --- | --- | --- | --- | --- |
| Labour | 347 | 103 (29.7) | 17.1 | 0.05 | 49.0 | 0.73 |
| Conservative | 116 | 0 | 23.2 | 0.04 | 48.8 | 0.58 |
| Liberal Dem | 65 | 0 | 22.1 | 0.05 | 48.8 | 0.51 |
| The Green Party | 4 | 0 | 19.5 | 0.10 | 49.3 | 0.53 |
| Reform UK | 5 | 3 (60.0) | 23.8 | 0.05 | 48.7 | 0.40 |

\*As measured by the Index of Multiple Deprivation SD = Standard deviation

The five areas which returned a Reform UK MP had the highest prevalence of 17 out of 20 health conditions: Asthma; COPD; Chronic kidney disease; Coronary heart disease; Dementia; Depression; Diabetes; Epilepsy; Heart Failure; Hypertension; Learning Disabilities; Obesity; Peripheral arterial disease; Rheumatoid arthritis; Schizophrenia, bipolar disorder and psychoses; Stroke or transient ischaemic attack (**Figure 1 and Table 2**). For example, Reform UK constituencies had an Asthma prevalence of 7.44% and a COPD prevalence of 2.85% compared with 6.58% and 1.99% for Labour areas. Reform UK areas had a CHD prevalence of 3.90% compared with 2.98% in Conservative areas, and a depression prevalence of 14.05% compared with 12.84% in Liberal Democrat areas.

**Figure 1:**
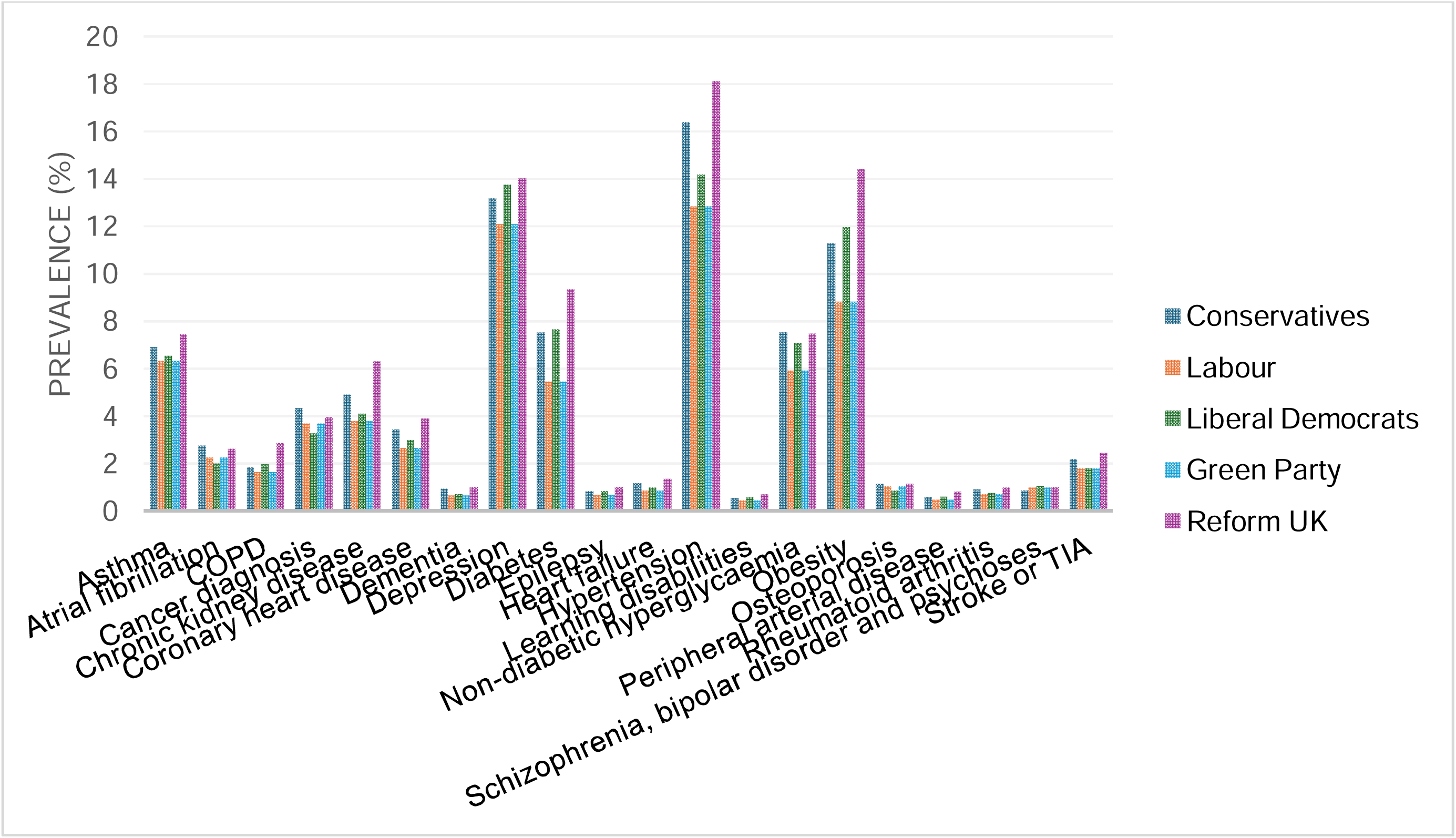
Prevalence of common conditions by winning party at 2024 General Election.

**Table 2:** prevalence of health conditions by winning party at 2024 UK General Election (highest prevalence in bold)

| **Condition** | **Winning party** | **Prevalence (%)** |
| --- | --- | --- |
| Asthma | Labour | 6.54 |
| Asthma | Conservative | 6.90 |
| Asthma | Liberal Dem | 6.84 |
| Asthma | Greens | 6.33 |
| **Asthma** | **Reform UK** | **7.44** |
| Atrial fibrillation (irregular heart rate) | Labour | 2.00 |
| Atrial fibrillation (irregular heart rate) | Conservative | 2.74 |
| **Atrial fibrillation (irregular heart rate)** | **Liberal Dem** | **2.65** |
| Atrial fibrillation (irregular heart rate) | Greens | 2.27 |
| Atrial fibrillation (irregular heart rate) | Reform UK | 2.59 |
| COPD | Labour | 1.97 |
| COPD | Conservative | 1.85 |
| COPD | Liberal Dem | 1.63 |
| COPD | Greens | 1.62 |
| **COPD** | **Reform UK** | **2.85** |
| Cancer diagnosis since 2003 | Labour | 3.27 |
| **Cancer diagnosis since 2003** | **Conservative** | **4.35** |
| Cancer diagnosis since 2003 | Liberal Dem | 4.31 |
| Cancer diagnosis since 2003 | Greens | 3.69 |
| Cancer diagnosis since 2003 | Reform UK | 3.95 |
| Chronic kidney disease | Labour | 4.10 |
| Chronic kidney disease | Conservative | 4.89 |
| Chronic kidney disease | Liberal Dem | 4.47 |
| Chronic kidney disease | Greens | 3.78 |
| Chronic kidney disease | Reform UK | 6.31 |
| Coronary heart disease | Labour | 2.98 |
| Coronary heart disease | Conservative | 3.41 |
| Coronary heart disease | Liberal Dem | 3.11 |
| Coronary heart disease | Greens | 2.65 |
| **Coronary heart disease** | **Reform UK** | **3.90** |
| Dementia | Labour | 0.70 |
| Dementia | Conservative | 0.92 |
| Dementia | Liberal Dem | 0.87 |
| Dementia | Greens | 0.66 |
| **Dementia** | **Reform UK** | **1.01** |
| Depression | Labour | 13.75 |
| Depression | Conservative | 13.16 |
| Depression | Liberal Dem | 12.84 |
| Depression | Greens | 12.10 |
| **Depression** | **Reform UK** | **14.05** |
| Diabetes | Labour | 7.65 |
| Diabetes | Conservative | 7.52 |
| Diabetes | Liberal Dem | 6.55 |
| Diabetes | Greens | 5.43 |
| **Diabetes** | **Reform UK** | **9.33** |
| Epilepsy | Labour | 0.83 |
| Epilepsy | Conservative | 0.81 |
| Epilepsy | Liberal Dem | 0.78 |
| Epilepsy | Greens | 0.67 |
| **Epilepsy** | **Reform UK** | **1.02** |
| Heart failure | Labour | 0.99 |
| Heart failure | Conservative | 1.16 |
| Heart failure | Liberal Dem | 1.02 |
| Heart failure | Greens | 0.85 |
| **Heart failure** | **Reform UK** | **1.34** |
| High blood pressure (hypertension) | Labour | 14.21 |
| High blood pressure (hypertension) | Conservative | 16.41 |
| High blood pressure (hypertension) | Liberal Dem | 15.02 |
| High blood pressure (hypertension) | Greens | 12.85 |
| **High blood pressure (hypertension)** | **Reform UK** | **18.15** |
| Learning disabilities | Labour | 0.59 |
| Learning disabilities | Conservative | 0.52 |
| Learning disabilities | Liberal Dem | 0.50 |
| Learning disabilities | Greens | 0.44 |
| **Learning disabilities** | **Reform UK** | **0.70** |
| Non-diabetic hyperglycaemia | Labour | 7.07 |
| **Non-diabetic hyperglycaemia** | **Conservative** | **7.56** |
| Non-diabetic hyperglycaemia | Liberal Dem | 7.23 |
| Non-diabetic hyperglycaemia | Greens | 5.92 |
| Non-diabetic hyperglycaemia | Reform UK | 7.46 |
| Obesity | Labour | 11.98 |
| Obesity | Conservative | 11.29 |
| Obesity | Liberal Dem | 9.92 |
| Obesity | Greens | 8.82 |
| **Obesity** | **Reform UK** | **14.40** |
| Osteoporosis | Labour | 0.86 |
| Osteoporosis | Conservative | 1.13 |
| **Osteoporosis** | **Liberal Dem** | **1.23** |
| Osteoporosis | Greens | 1.05 |
| Osteoporosis | Reform UK | 1.14 |
| Peripheral arterial disease | Labour | 0.60 |
| Peripheral arterial disease | Conservative | 0.59 |
| Peripheral arterial disease | Liberal Dem | 0.55 |
| Peripheral arterial disease | Greens | 0.47 |
| **Peripheral arterial disease** | **Reform UK** | **0.79** |
| Rheumatoid arthritis | Labour | 0.75 |
| Rheumatoid arthritis | Conservative | 0.89 |
| Rheumatoid arthritis | Liberal Dem | 0.79 |
| Rheumatoid arthritis | Greens | 0.71 |
| **Rheumatoid arthritis** | **Reform UK** | **0.98** |
| Schizophrenia, bipolar disorder and psychoses | Labour | 1.05 |
| Schizophrenia, bipolar disorder and psychoses | Conservative | 0.84 |
| Schizophrenia, bipolar disorder and psychoses | Liberal Dem | 0.87 |
| Schizophrenia, bipolar disorder and psychoses | Greens | 0.97 |
| **Schizophrenia, bipolar disorder and psychoses** | **Reform UK** | **1.03** |
| Stroke or transient ischaemic attack | Labour | 1.81 |
| Stroke or transient ischaemic attack | Conservative | 2.17 |
| Stroke or transient ischaemic attack | Liberal Dem | 2.07 |
| Stroke or transient ischaemic attack | Greens | 1.80 |
| **Stroke or transient ischaemic attack** | **Reform UK** | **2.43** |

Vote share for Reform UK in individual constituencies ranged from 0 to 46.2%, with positive correlations between Reform UK vote share and prevalence for 18/20 health measures (**Table 3 and Figure 2**) Seven of these were strong correlations (>0.5) and 10 were moderately sized (i.e. between 0.3 and 0.49). The strongest correlations were for obesity (0.654) and COPD (0.647) and epilepsy (0.632) all p<0.001.

**Figure 2.**
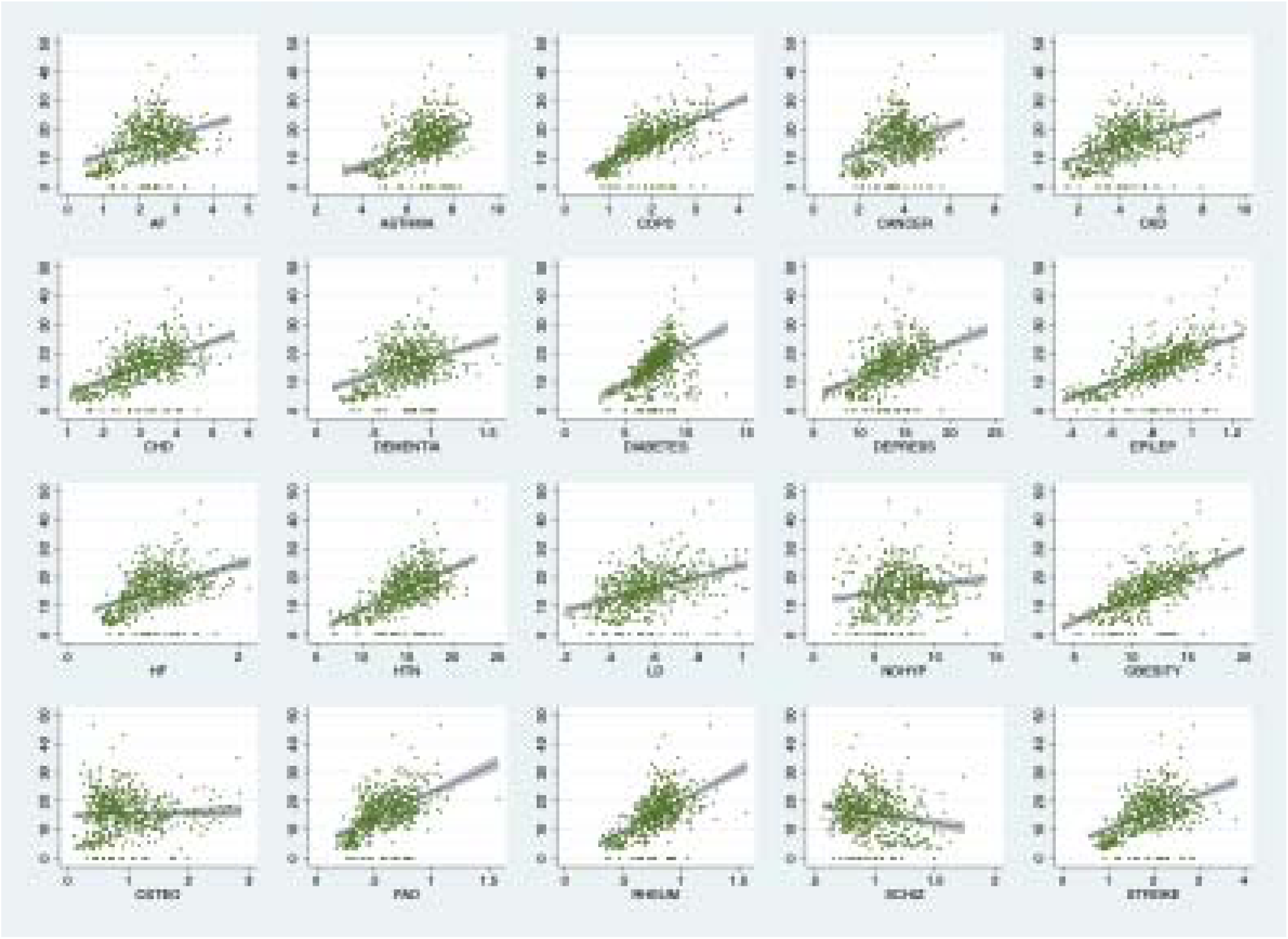
Correlations between Reform UK vote proportions and selected non-communicable diseases

**Table 3:** Unadjusted Pearson correlation coefficients between Reform UK vote share and health outcomes at the constituency level in England. Sorted largest to smallest.

| Condition | Correlation | p-value |
| --- | --- | --- |
| Obesity | 0.654 | <0.001 |
| COPD | 0.647 | <0.001 |
| Epilepsy | 0.632 | <0.001 |
| Hypertension | 0.577 | <0.001 |
| Rheumatoid arthritis | 0.548 | <0.001 |
| Coronary Heart Disease | 0.537 | <0.001 |
| Peripheral Arterial Disease | 0.520 | <0.001 |
| Depression | 0.479 | <0.001 |
| Asthma | 0.471 | <0.001 |
| Diabetes | 0.468 | <0.001 |
| Stroke or TIA | 0.457 | <0.001 |
| Chronic Kidney Disease | 0.448 | <0.001 |
| Heart Failure | 0.442 | <0.001 |
| Learning Disability | 0.405 | <0.001 |
| Dementia | 0.388 | <0.001 |
| Atrial fibrillation | 0.382 | <0.001 |
| Cancer | 0.323 | <0.001 |
| Non-diabetic hyperglycaemia | 0.174 | <0.001 |
| Osteoporosis | 0.036 | 0.3988 |
| Schizophrenia, bipolar disorder and psychoses | -0.187 | <0.001 |

**Table 4:** Relationship between Reform UK vote share and prevalence of health conditions.

| Condition | Coeff | p value | Lower CI | Upper CI |
| --- | --- | --- | --- | --- |
| **Asthma** | 0.113 | 0.011 | 0.026 | 0.201 |
| **Atrial fibrillation** | 0.026 | 0.061 | -0.001 | 0.053 |
| **Cancer** | 0.029 | 0.190 | -0.014 | 0.071 |
| **Chronic Kidney Disease** | 0.251 | <0.001 | 0.120 | 0.381 |
| **COPD** | 0.261 | <0.001 | 0.213 | 0.309 |
| **Coronary Heart Disease** | 0.119 | <0.001 | 0.072 | 0.166 |
| **Dementia** | 0.023 | 0.002 | 0.008 | 0.037 |
| **Depression** | 0.980 | <0.001 | 0.654 | 1.306 |
| **Diabetes** | 0.459 | <0.001 | 0.305 | 0.614 |
| **Epilepsy** | 0.068 | <0.001 | 0.054 | 0.082 |
| **Heart Failure** | 0.030 | 0.039 | 0.002 | 0.059 |
| **Hypertension** | 0.824 | <0.001 | 0.677 | 0.970 |
| **Learning Disability** | 0.025 | 0.002 | 0.009 | 0.041 |
| **Non-diabetic hyperglycaemia** | -0.098 | 0.523 | -0.400 | 0.204 |
| **Obesity** | 1.479 | <0.001 | 1.239 | 1.720 |
| **Osteoporosis** | -0.056 | 0.109 | -0.125 | 0.013 |
| **Peripheral Arterial Disease** | 0.027 | 0.001 | 0.011 | 0.043 |
| **Rheumatoid arthritis** | 0.057 | <0.001 | 0.043 | 0.072 |
| **Schizophrenia, bipolar disorder and psychoses** | -0.093 | <0.001 | -0.113 | -0.073 |
| **Stroke or TIA** | 0.019 | 0.155 | -0.007 | 0.046 |

Results from separate linear regressions, adjusted for % of local area over the age of 65 years; sex; and IMD in five groups. Results shown for Reform UK vote share only and represent changes in percentage of population with health conditions per 10% increase in Reform UK vote share.
CI = confidence interval

After controlling for age, sex and IMD, there were statistically significant positive relationships between Reform UK vote share and prevalence of 15 /20 conditions studied at p<0.05 (**Table** 4)he largest association was for obesity where a 10% increase in Reform UK vote share was associated with a +1.479% increase in obesity prevalence (95% Confidence Interval 1.239 to 1.720). For each 10% increase in Reform UK vote share there was a +0.261% (95%CI 0.213 to 0.309) higher prevalence of COPD, a +0.113(0.026 to 0.201)% prevalence of asthma, and a +0.980(0.654 to 1.306)% prevalence of depression.

## DISCUSSION

The main finding of the present analysis was an association between poor health metrics at a constituency level and votes for Reform UK. The five constituencies where the party won the seat had the highest prevalence for 15 out of 20 health conditions, and across the country there were positive correlations between the percentage vote for Reform UK and the constituency prevalence for 18 out of 20 of them. These relationships persisted when adjusted for age, sex and level of deprivation level of constituencies.

The results findings are consistent with work showing a relationship between poor healthcare measures and Republican voting in the US[5] and data from Italy linking dissatisfaction with public services and voting for the far right[6]. In the UK, closure of local healthcare facilities has been shown to reduce reported patient satisfaction and increase support for populist right parties[7].

Lung health is particularly influenced by health inequality[1-4], and conditions causing breathlessness (obesity, COPD as well as asthma and cardiac disease) are linked to voting pattern. The choice to pursue austerity policies, aggravated by the effects of the COVID-19 pandemic, have contributed to the fact that most people with long-term lung conditions are missing out on basic aspects of care, which may fuel frustration with the status quo[1 2 4]. Living in a home that is cold and or damp is associated with an increased risk of acute exacerbations and hospitalisation, so poor housing will also interact with health experience[4].

Three of the five Reform voting constituencies are coastal. The 2021 Chief Medical Officer’s report[13] highlighted the particular health issues there, due to a combination of greater ill health in older poorer populations with more long term conditions, while healthcare provision is lower than other parts of the country, a specific manifestation of the inverse care law[14].

Depression can be characterised by negative feelings about the self, world, and future[15], as well as a reduction in the sense of being in control of one’s life[16 17] and a recent US study has found an association between depression and misperception as to the legitimacy of election results[18]. Experience of mental as well as physical health issues may therefore influence decisions around voting.

## Limitations

We acknowledge some limitations to the present study. Election data come from 2024 while health indicators come from 2022/23 (the latest available at these geographies). Both health and political sentiment are influenced by long term trends which are not captured in this cross-sectional study. Approximately 90% of constituency boundaries were changed for the 2024 election, so longitudinal analyses are difficult to conduct. Finally, as analyses are at aggregate geographies, the ecological fallacy is possible here and only limited conclusions about individual behaviour can be drawn from average population characteristics. Future work, both quantitative and qualitative, should address these issues and potential mechanisms.

## Conclusion

Government holds many levers that can influence health, including decisions around spending on healthcare and other aspects of infrastructure such as transport and education, redistributive policies to reduce poverty, legislation to address commercial determinants of health, as well as social and community services to support people living with long term health conditions[3]. While these analyses are ecological, they do suggest poorer health as linked to increased votes for Reform UK and suggest a useful synergy across the political spectrum. For Reform UK policy makers, they demonstrate that there are profound health issues in their constituencies which should be addressed.For those elsewhere in the political spectrum, they should provide a further incentive to take steps to improve public health and reduce health inequalities.

## DATA SHARING

Datasets used for this research are publicly available.

## AUTHOR CONTRIBUTION

NSH and AAL designed the study. AAL conducted the analysis. AAL and NSH produced the first draft and edited this. All authors have reviewed and approved the final manuscript and AAL is the guarantor.

## FUNDING

The funders had no input into data analysis or the writing of this manuscript.

## TRANSPARENCY DECLARATION

Anthony Laverty, the manuscript’s guarantor affirms that the manuscript is an honest, accurate, and transparent account of the study being reported; that no important aspects of the study have been omitted; and that any discrepancies from the study as planned (and, if relevant, registered) have been explained.

## Data Availability

All data are available online:
House of Commons Library data and other sources listed below were all accessed 22/11/2024.
General election 2024 results:
https://commonslibrary.parliament.uk/research-briefings/cbp-10009/
https://www.parliament.uk/about/how/elections-and-voting/constituencies/
Constituency data: Population, by age
https://commonslibrary.parliament.uk/constituency-statistics-population-by-age/
Index of multiple deprivation by constituency
https://commonslibrary.parliament.uk/constituency-data-how-healthy-is-your-area/
Quality and Outcomes Framework Data
https://github.com/houseofcommonslibrary/local-health-data-from-QOF

## Notes

### Competing Interest Statement

NSH is Chair of Action on Smoking and Health and Medical Director of Asthma and Lung UK. AAL is a Trustee of Action on Smoking and Health.

### Funding Statement

This study did not receive any funding

